# Clinical prediction models for treatment outcome in newly-diagnosed epilepsy: Protocol for a systematic review

**DOI:** 10.1101/2022.05.05.22274710

**Authors:** Corey Ratcliffe, Anthony Marson, Simon S. Keller, Laura J. Bonnett

## Abstract

Epilepsy, characterised by a predisposition towards unprovoked seizures, is one of the most common neurological disorders globally. Whilst 60-70% of individuals diagnosed with epilepsy will gain seizure control through anti-seizure medication, the mechanisms underlying seizure persistence are unclear. Intractability can significantly degrade a patient’s quality of life amongst other things; the use of predictive modelling of epilepsy outcomes in deciding on treatment therefore offers a tangible benefit to the patient. Early indicators of pharmacoresistance may discourage certain treatment options, and save time in what has been indicated to be a critical stage for newly-diagnosed epilepsy. Primarily, this paper aims to evaluate existing predictive models to identify demographic, clinical, physiological (e.g. EEG), and neuroimaging (e.g. MRI) factors that may be predictive of treatment outcomes in newly-diagnosed epilepsy. Two electronic databases, MEDLINE and EMBASE, will be searched with terms related to prognosis in newly-diagnosed epilepsy, and identified studies will be included for review if they have combined at least two demographic, clinical, neuroimaging, and/or physiological factors to predict treatment outcome in people with newly-diagnosed epilepsy. Papers will be screened by two independent reviewers via titles, abstracts and then full text against the inclusion criteria for eligibility. Data will be extracted by reviewers using standardised forms, assessed for risk of bias using the PROBAST tool and synthesised narratively. If considered appropriate the authors will carry out a meta-analysis on the available data.

**Prospero registration number:** – CRD42022329936

## Background

### Description of the condition

Epilepsy is a neurological disorder that is estimated to affect 50-70 million people worldwide, defined as a predisposition towards generating unprovoked seizures.^1,2^ Epilepsy is more common in developing nations despite not being dependent on race, more common in males although only marginally so, and poses a higher risk to the elderly or those under one year of age.^3^ Diagnosis follows two unprovoked seizures or one seizure with a high risk (at least 60%) of recurrence over the next 10 years.^4^ Epilepsy is not a single disorder, but encompasses numerous syndromes as defined by the International League Against Epilepsy (ILAE).^5^

At diagnosis the standard clinical practice for treatment is monotherapy, where the appropriate anti-seizure medication (ASM) is selected based on numerous factors.^6,7^ However, 30-40% of patients will not achieve seizure remission, and if seizures persist subsequent to two ASM trials (either monotherapy or polytherapy), the patient will be considered pharmacoresistant.^8–10^ Seizure persistence has been correlated with psychological, psychiatric, cognitive, and social comorbidities.^11^

### Rationale for the review

Whilst there are patients for whom treatment can be safely withheld, the prognosis for newly-diagnosed epilepsy (NdE; a diagnosis of epilepsy no more than three months prior) is typically optimal following timely intervention (i.e. after a minimal number of seizures).^6,11–13^ In many cases of NdE, the first-line ASM may be retained as the long-term monotherapy, however, in 40-50% of patients this is not sufficient to gain seizure control.^14–16^ Depending on certain prognostic factors (i.e. any health-outcome associated factor in an individual with a specific health condition) adjunctive therapy (polytherapy) may be more appropriate; and indeed, in pharmacoresistant focal epilepsy, resective surgery has a desirable outcome.^17–19^ Through better characterisation of early prognostic factors, patients and clinicians can expect clearer estimates of treatment outcomes, ultimately leading to more informed treatment decisions and higher remission rates.

To date, there have been numerous investigations of prognostic factors of treatment outcome in epilepsy.^11,20–24^ Where the 2014 review by Abimbola et al. offers the most recent systematic approach, this current review will examine predictive models to not only identify prognostic factors, but also evaluate the models’ performance and risk of bias; which will be assessed using the recently published PROBAST tool, addressing any concerns about applicability.^25^ Furthermore, the search criteria for this review will also include studies with sample sizes of fewer than 100, and studies published subsequent to the aforementioned investigations.

### Research aims

This study aims to systematically review the available evidence for prediction models of treatment outcome(s) in NdE. Our primary objective is to identify the factors that are significantly associated with 12 months continuous freedom from seizures.

### Methods Selection criteria

#### Study design

This review will include published reports of any study design (including, but not limited to: cohort studies, randomised control trials, quasi-randomised control trials, observational studies, and case-control studies) that use prediction models to prognosticate treatment response in NdE.

#### Participants

This review will include studies involving any person with NdE defined using the operational ILAE definition of two clinically unprovoked seizures, or one unprovoked seizure with a ≥ 60% probability of recurrence.^4^ Studies using an alternative definition of newly-diagnosed epilepsy will be assessed for inclusion on a case by case basis according to their agreement with the ILAE definition.

#### Prediction models

This review will identify prediction models developed with at least two demographic, and/or clinical neuroimaging, and/or electrophysiological factors collected and assessed as part of standard clinical practice upon a new diagnosis of epilepsy that are associated with 12-month seizure remission. Demographic factors are socioeconomic attributes that can be statistically expressed—for example age, sex, and education level. Clinical factors are signs and symptoms of disease classification or severity including aetiology, type and frequency of seizure, age of onset of epilepsy and duration of illness prior to diagnosis. The neuroimaging and neurophysiological factors include assessments of standard MRI and EEG examinations respectively, often taken upon the new diagnosis of epilepsy.

### Outcome measures

#### Primary outcome

- 12-month / 48-week remission

ILAE reports suggest that 48 weeks (or 12 months) is the ideal time at which to evaluate the long-term efficacy of treatment.^26,27^ Alongside clinical relevance, 12 months has ecological importance to individuals diagnosed with epilepsy; for example, 12 months without seizure is the requirement by the DVLA to allow a patient with epilepsy to drive.^28^ This study will therefore include prediction models of 12-month remission (freedom from seizures).

#### Secondary outcomes

- Remission (of any duration) at any reported time point
- Treatment failure (adverse effects, intractability, etc.), reported in any form and at any time

### Search methods for identification of studies

#### Electronic searches

Broad electronic searches will be conducted using MeSH headings and free text words/phrases with no language or date restrictions in the following databases: MEDLINE, EMBASE. These searches will include terms related to epilepsy, clinical and neuroimaging factors, treatment outcomes, and prediction models—the latter of which will be targeted using the Ingui prediction modelling search strategy.^29^

#### Search strategy

The search strategy has been developed in consultation with a medical librarian experienced in literature searching for systematic reviews. The strategy is included in Table 1.

**Table 1.** The search strategy used for database investigation.

| # | Searches |
| --- | --- |
| 1. | early diagnosis/ |
| 2. | ((recent\$ OR new\$ OR early) adj2 (diagnos\$ OR onset)).tw. |
| 3. | ("first seizure" OR "first fit").tw. |
| 4. | 1 OR 2 OR 3 |
| 5. | exp Epilepsy/ OR epilep\$.tw. |
| 6. | (validation studies OR clinical trial OR clinical trial phase i OR clinical trial phase ii OR clinical trial phase iii OR clinical trial phase iv OR comparative study OR evaluation studies OR multicenter study).pt. |
| 7. | ((observation\$ OR cohort OR case\$ OR cross?section\$ OR "cross section\$" OR "time-series" OR "time series" OR "before and after" OR "before-and-after" OR retrospective) adj2 (study OR trial OR method)).mp. |
| 8. | (randomized controlled trial OR controlled clinical trial).pt. OR (randomized OR placebo OR randomly).ab. |
| 9. | clinical trials as topic.sh. |
| 10. | trial.ti. |
| 11. | 6 OR 7 OR 8 OR 9 OR 10 |
| 12. | exp animals/ not humans.sh. |
| 13. | 11 not 12 |
| 14. | 13 not case reports.pt. |
| 15. | (Validat\$ OR Predict\$.ti. OR Rule\$) OR (Predict\$ AND (Outcome\$ OR Risk\$ OR Model\$)) OR ((History OR Variable\$ OR Criteria OR Scor\$ OR Characteristic\$ OR Finding\$ OR Factor\$) AND (Predict\$ OR Model\$ OR Decision\$ OR Identif\$ OR Prognos\$)) OR (Decision\$ AND (Model\$ OR Clinical\$ OR Logistic Models/)) OR (Prognostic AND (History OR Variable\$ OR Criteria OR Scor\$ OR Characteristic\$ OR Finding\$ OR Factor\$ OR Model\$)) |
| 16. | 5 AND 14 AND 15 |
| 17. | 4 AND 16 |

#### Searching other resources

The search will be supplemented by identifying potentially relevant studies from other sources, such as searching the references of relevant existing systematic reviews of prognosis in NdE, and the reference lists of all included studies. Any prediction models identified via validation investigations will also be considered in place of the validation study.

### Data collection and analysis

#### Selection of studies

Independent screening of all citations identified by the search will be carried out by two review authors (CR and LB). At this stage papers will be screened by their titles and abstracts according to the inclusion criteria, and publications that do not meet this will be excluded. Citations will then be screened by full-text assessment for eligibility. The two reviewers will discuss any inconsistencies found in their screening and if consensus cannot be reached, a third reviewer (SK) shall be consulted to resolve them.

#### Data extraction and management

Two independent reviewers (CR and LB) will conduct data extraction via pre-piloted data extraction forms. Any discrepancies between reviewers will be resolved through discussion and, should a consensus not be reached, a third reviewer (SK) will be consulted for judgment. If data is missing or additional data is required, the reviewers will attempt to contact the corresponding authors of studies in order to obtain it. The data extracted from studies for this review will include:

- Details of study characteristics (e.g. date of study, country in which the study takes place, etc.)
- Study design characteristics (e.g. type of trial, prospective or retrospective study, etc.)
- Patient demographics (e.g. age, gender, treatment, etc.)
- Outcome measures (prediction models of seizure freedom, rate of treatment failure)

#### Assessment of methodological quality

Two independent reviewers (CR and LB) will assess the quality of individual studies in accordance with the Prediction model Risk Of Bias ASsessment Tool (PROBAST), which addresses four potential domains of bias: predictors, participants, outcome, and analysis.^25^ Potential disagreements will be solved through discussion or the judgment of a third review author.

The PROBAST results for each included study will be presented in tabular format—both by domain and overall. Reviewers will first assess the relevant risk of bias of items in each domain and then produce an overall judgement based on these ratings. Each domain and overall risk of bias will be categorised as low, moderate or high risk of bias. An overall judgement of low risk of bias will require all four bias domains to be rated as low risk of bias.

#### Data synthesis

Data pertinent to describing the setting, demographics, primary outcomes, and quality assessment for individual studies will be synthesised in narrative form and in evidence tables. Following this, if of suitable quality and volume, meta-analysis may be considered according to the methodology prescribed by Debray et al.^30^

#### Analysis of subgroups

If sufficient data is available, the review may separate focal and generalised epilepsies for subgroup analysis.

## Discussion

It is hypothesised that early treatment with a single therapeutic agent will offer the best prospect of achieving seizure-freedom in NdE.^6^ As such, developing information on early diagnosis and selecting the most appropriate treatment for a patient with NdE is of significant importance. This review therefore intends to summarise existing prediction models for treatment outcomes in people with NdE. The authors hope this review will contribute to expediting the treatment selection process, both by collating existing information regarding factors that prognosticate treatment outcomes, and by identifying any as-yet-underpowered clinical, demographic, neurophysiological, or neuroimaging factors presented in the literature.

## Data Availability

All data produced in the present study are available upon reasonable request to the authors.

